# Knowledge and attitudes toward infertility and reproductive health among nursing and midwifery students from selected universities in Accra, Ghana

**DOI:** 10.64898/2026.11.06.26367415

**Authors:** Ernestina Safoa Donkor, Emmanuel Kojo Osafo, Ezekiel Oti-Boadi, Benedicta Twum-Dei

**Affiliations:** Department of Midwifery, School of Nursing and Midwifery, Heritage Christian University, Amasaman, Accra, Ghana; Last Mile Health and Bitblue Consult, Accra, Ghana; Department of Nursing, School of Nursing and Midwifery, Heritage Christian University, Amasaman, Accra, Ghana; Department of Nutrition and Dietetics, School of Allied Health Sciences, Heritage Christian University, Amasaman, Accra, Ghana

**Keywords:** Ghana, Infertility, Knowledge-attitude-practice, Nursing education, Nursing/midwifery students, Reproductive health

## Abstract

**Introduction:** In sub-Saharan Africa, where high societal value is placed on childbearing, infertility prevalence is high and often leads to stigma and psychosocial distress. Nurses and midwives, as frontline reproductive health providers, need adequate preparation to support infertile couples, yet a literature gap exists regarding how well nursing and midwifery trainees are equipped with the knowledge and attitudes needed for this role. This study investigated the knowledge and attitudes regarding infertility among nursing and midwifery students, applying the Knowledge-Attitude-Practice (KAP) model.

**Materials and methods:** A descriptive cross-sectional survey was conducted among a stratified sample of 263 final-year BSc nursing and midwifery students at four universities in Accra, Ghana. Data were collected via a structured questionnaire covering demographics, infertility knowledge, attitudes, and practice intentions; and were analysed using SPSS.

**Results:** Respondents demonstrated a high overall level of infertility knowledge (composite knowledge score: M = 18.25/23, SD = 3.17; attitudes toward infertility were largely favourable (composite attitude score: M = 3.812/5, SD = 0.654; female respondents scored significantly higher than males on both knowledge (U = 2800.5, p = .044) and attitude (U = 2777.0, p = .040). BSc Midwifery students demonstrated significantly higher knowledge than BSc Nursing students (U = 2868.0, p = .002). A statistically significant moderate positive correlation was found between knowledge and attitude scores (r = .400, p < .001), and knowledge score was the sole significant predictor of attitude in multiple regression (B = 0.080, p < .001, R² = .178). The large majority of respondents (86.3%) supported expanded infertility content in the curriculum.

**Conclusion:** The significant association between knowledge and attitude supports the KAP framework and suggests that targeted curricular reinforcement of weak knowledge areas may further strengthen students’ attitudes. The findings will inform curriculum improvements and promote evidence-based practice in nursing and midwifery education.

## Introduction

Infertility, defined clinically as the failure to achieve a pregnancy after 12 months or more of regular unprotected sexual intercourse, is recognized by the World Health Organization (WHO) as a disease of the reproductive system [1]. Globally, infertility affects one in six individuals, with this prevalence strikingly consistent across high-, middle-, and low-income countries (around 16-18%), underscoring that infertility is a universal public health concern [1]. The burden of infertility is particularly acute in sub-Saharan Africa, including Ghana, due to strong pronatalist cultural values. In many African societies, childbearing is not only deeply desired but also socially expected as a fulfilment of marriage and a source of support and status [2, 3]. Consequently, couples who cannot have children may face intense societal pressure and stigma. In Ghana, epidemiological estimates suggest that roughly 11-12% of women and 15-16% of men of reproductive age experience infertility [4]. Notably, male-factor infertility is a contributing cause in a substantial proportion of cases; however, men are often excluded from the infertility discourse in traditional settings, with blame and social repercussions disproportionately falling on women [3, 5]. This gendered attribution means Ghanaian women suffering infertility can be stigmatized as “barren” and may endure various forms of discrimination and abuse.

Within the socio-cultural context of Ghana, the consequences of infertility for women are severe. Qualitative studies have documented that women struggling to conceive often experience psychological trauma and social ostracism. For example, a recent study in urban Ghana found that infertile women frequently reported loss of self-esteem, marital instability, social isolation, and instances of physical and emotional abuse as a result of their condition [6]. Such women may be pressured by spouses or in-laws, sometimes leading to polygamy or divorce as the husband seeks children with another partner [6, 7]. Infertility is thus not only a medical issue but also a profound social challenge in Ghana. Many affected couples turn to informal or traditional solutions before seeking biomedical care. Studies have noted that some Ghanaians attribute infertility to spiritual causes or past transgressions, and herbal remedies and spiritual interventions are commonly sought [5]. These beliefs can delay timely medical consultation and contribute to misconceptions about infertility and its treatments.

Healthcare professionals, especially nurses and midwives, have a crucial role in addressing infertility and mitigating its impacts. In Ghana’s healthcare system, nurses and midwives are often the first point of contact for women’s health issues, including family planning and antenatal care. However, there appears to be a gap in both research and education regarding infertility in the nursing/midwifery curriculum. Globally, reproductive health training for nurses tends to focus on maternal health and contraception, often giving limited attention to infertility and subfertility care [8]. In Ghana, issues of infertility have historically been under-prioritized in public health initiatives compared to problems like high fertility and maternal mortality [9]. As a result, future nurses/midwives may be under-prepared to handle infertility-related cases or to educate the community.

Their position in communities and clinics means nurses and midwives are well-placed to provide education, counselling, and support to couples with fertility problems. Competent, empathetic nursing care can help dispel myths, encourage early referral to specialists, and provide much-needed psychosocial support to infertile individuals [10]. Conversely, if nurses and midwives themselves lack adequate knowledge or hold prejudiced attitudes about infertility, they may unknowingly reinforce stigma or fail to offer appropriate guidance. Furthermore, if future nurses and midwives hold misconceptions or negative biases, for example believing infertility is always the woman’s “fault” or that assisted reproductive technologies are morally unacceptable, such attitudes could adversely affect the quality of care they provide to individuals and couples facing infertility. It is therefore vital that nursing and midwifery education prepares students with sound knowledge of infertility, its causes, prevention, and treatments, and fosters positive, nonjudgmental attitudes toward those affected. Addressing this problem is essential for improving reproductive healthcare services and reducing infertility-related stigma.

The problem that guided this study is the unknown level of knowledge and the nature of attitudes toward infertility among nursing and midwifery students in Ghana. Despite being key stakeholders in reproductive healthcare delivery, student nurses and midwives may lack the knowledge and empathetic orientation necessary to support infertile individuals, and there is insufficient empirical evidence on their preparedness in this regard. A preliminary review of the literature reveals a paucity of empirical research concerning Ghanaian nursing and midwifery students’ understanding of infertility. Most existing studies in the region have centered on either general population awareness [11] or the experiences of infertile women [5, 6, 12], rather than on the competencies of healthcare trainees. Without knowing the knowledge gaps and attitudes of student nurses and midwives, educational institutions cannot effectively tailor their curricula or interventions to address those gaps. This research is significant because it addresses this gap in the literature by providing empirical data on the preparedness of nursing and midwifery students to deal with infertility, and the findings can inform educators and policymakers about critical content areas that may need strengthening in the nursing and midwifery curricula.

The aim of this study was therefore to assess the knowledge and attitudes regarding infertility among BSc final-year (Level 400) nursing and midwifery students in four (4) tertiary institutions in Accra, Ghana. The specific objectives of the research were to: (i) assess the level of knowledge about infertility among final-year nursing and midwifery students; (ii) determine the attitudes of final-year nursing and midwifery students toward infertility; (iii) determine how demographic factors relate to knowledge and attitude; and (iv) ascertain the association between students’ knowledge level and their attitudes.

This study was anchored in the Knowledge-Attitude-Practice (KAP) model, a theoretical framework frequently used in public health and health education research. The KAP model posits a sequential and interrelated progression: individuals first acquire knowledge about a topic, which shapes their attitudes or beliefs, and in turn influences their practices or behaviours; improved knowledge is theorized to lead to more positive attitudes and ultimately to better practices [13]. The model has been widely applied through KAP surveys, structured questionnaires designed to quantitatively assess what respondents know, how they feel, and how they behave with respect to a specific issue [14]. An advantage of the KAP model is that it provides a systematic approach for identifying knowledge gaps, misconceptions, and attitudinal barriers that may hinder effective practice [15]. In the context of infertility, a KAP-based study can reveal whether nursing and midwifery students know key facts, such as the definition of infertility or its common causes, whether they hold biases or stigma toward infertile people, and whether they intend to apply best practices such as making appropriate referrals or offering counselling in the future.

The KAP framework assumes a rational linkage between knowledge gain and behaviour change, a notion that has been both utilized and critiqued in health education literature. While increased knowledge alone does not always guarantee attitude change or improved practice, it is generally a prerequisite for informed action. In this study, the KAP model guided the development of the survey instrument and the interpretation of results. Knowledge items measured the cognitive domain, whereas attitude items gauged the affective domain. Although practice in the strict sense could not be fully assessed in a student population not yet independently practicing, the survey included items on intended practices and self-reported behaviours. By using the KAP model, the study could not only describe knowledge and attitudes but also explored the relationship between these components, for instance whether students with higher knowledge scores also exhibited more positive attitudes toward infertility, aligning with similar studies that have employed this model to investigate health professionals’ perspectives on reproductive health topics [16, 17].

## Materials and methods

### Study design

This research employed a descriptive cross-sectional survey design. A cross-sectional approach was appropriate because the aim was to capture a “snapshot” of the knowledge and attitudes of nursing and midwifery students at a specific point in time.

### Population and sampling

The research was conducted in Greater Accra, Ghana, at four universities offering BSc nursing and midwifery programmes These institutions were selected to represent public and private universities that train nurses and midwives, thereby providing a diverse student sample within the region. The survey was administered after obtaining permission from each institution’s administration. By covering multiple institutions, the study design enhanced the generalizability of results to similar educational contexts in Ghana, rather than being confined to a single school’s culture or curriculum. Accra, as Ghana’s national capital, hosts a high concentration of accredited nursing and midwifery institutions spanning public and private ownership, faith-based and secular affiliations, and both generalist and specialist healthcare orientations, providing a diverse and meaningful sample frame even without national geographic dispersion.

The target population comprised all final-year (Level 400) nursing and midwifery students enrolled at the four selected universities. This cohort was targeted because they had completed most of their nursing/midwifery coursework and clinical rotations, including any content on reproductive health, and were therefore expected to have the highest level of knowledge and more formed attitudes, with their views indicative of what they would carry into the workforce imminently.

Using a stratified approach where each institution serving as a distinct stratum, all Level 400 nursing and midwifery students were eligible. A total of 263 students consented and participated.

### Measures

Data were collected using a structured self-administered questionnaire developed by the researchers based on the KAP framework and informed by the literature on infertility knowledge and attitudes [11, 16, 17]. The instrument comprised four sections. Section A (Demographics) gathered background information including age, sex, marital status, programme, year of study, and any personal or family experience with infertility. Section B (Knowledge of Infertility) used a mix of question formats to assess factual knowledge concerning definition and duration, prevalence, treatment, and prevention. Section C (Attitudes toward Infertility) measured attitudes using Likert-scale statements derived from previous attitude scales and tailored to the local context, with respondents indicating their level of agreement on a 5-point Likert scale (1 = Strongly Disagree, 5 = Strongly Agree). Section D (Practices/Intentions) captured relevant behaviour or intended behaviour, including whether respondents had received formal teaching on infertility, whether they had encountered infertility cases during clinical rotations, their likelihood of participating in future continuing education on fertility, and their perceived role in infertility prevention.

Content validity of the instrument was established through expert review. The draft questionnaire was given to three experts: a senior nursing lecturer knowledgeable in reproductive health, a gynaecologist, and a public health expert, who reviewed the items for relevance, clarity, and cultural appropriateness. Their feedback led to modifications such as rewording ambiguous questions and adding items, including an item on the emotional impact of infertility to reflect empathy. A pre-test (pilot) was conducted with nine (9) nursing students from a different university, not among the four (4) study sites, to check for any difficulties in understanding the questions; pilot respondents completed the survey and were interviewed about any confusing items, and minor adjustments were made accordingly. To ensure reliability, particularly of the attitude scale, Cronbach’s alpha for the attitude items from the pilot data was calculated to be .739 and the main study was .850.

#### Data collection procedure

After ethical clearance and administrative approvals, the researchers coordinated with faculty and class representatives at each university. Primary data were collected electronically using a self-administered online survey hosted on Google Forms. To reach the entire population of Level 400 nursing and midwifery students efficiently across the four (4) institutions, the survey URL was disseminated through official class communication platforms, specifically class WhatsApp groups, with the help of the respective class representatives. The landing page of the Google Form contained a mandatory digital informed consent form outlining the study’s purpose, anonymity clauses, and voluntary nature. Participants could only access the questionnaire sections after explicitly consenting to participate. To enforce data integrity and prevent duplicate entries, the platform configuration was restricted to one response per user via Google’s authentication control. Data collection remained open for a period between Monday 3^rd^ November and Wednesday 31^st^ December 2025, during which periodic follow-up reminders were posted to maximize the response rate.

### Data analysis

Data analysis was performed using Statistical Package for the Social Sciences (SPSS), version 26. Both descriptive and inferential statistics were utilized. Frequencies and percentages were computed for categorical variables, and means with standard deviations for continuous variables. Individual knowledge items were analysed for the percentage of students answering correctly. A composite knowledge score was computed by assigning 1 point for each correct response and 0 for incorrect or “not sure” responses across all knowledge items; for multiple-response items (B3, B4, B8), 1 point was awarded per scientifically valid option selected, and 1 point deducted for clearly inaccurate responses such as spiritual causes, with “Other, specify” responses reviewed and scored for scientific validity. The maximum total knowledge score was 23, and respondents were categorized into Poor (0-7), Moderate (8-15), and Good (16-23) knowledge levels, following similar scoring models used in reproductive health KAP studies (Mabitsela et al., 2025b). For attitude items, the distribution of agreement responses was examined for each statement, and overall attitude was summarized by the mean item score, ranging from 1 to 5.

Bivariate analyses were conducted to compare knowledge and attitude by key demographic factors. The relationship between knowledge and attitudes was assessed using Pearson’s correlation coefficient. Chi-square tests were applied to relevant categorical cross-tabulations, and where multiple factors appeared to influence knowledge or attitude, multiple regression analysis was used. All statistical tests were conducted at a 95% confidence level, with a p-value of < .05 considered statistically significant.

### Ethical considerations

This study was reviewed and approved by the Dodowa Health Research Centre Institutional Review Board (DHRCIRB), which served as the primary ethics committee given the lead researchers’ affiliation with that institution. Official permission letters were also obtained from the authorities of the four participating universities to allow data collection among their students. Participation was entirely voluntary, and informed consent was obtained from all participants. Students were informed that choosing not to participate would have no consequences on their academic standing, and only those who provided informed consent proceeded to fill out the questionnaire. Students could skip any question they were not comfortable with, or withdraw from the survey at any point before submission. Measures were taken to ensure that responses could not be traced back to an individual; the questionnaires did not collect names or student IDs, and all data were handled in aggregate form, with institution names coded rather than explicitly named in results to avoid stigma or unwarranted comparisons. Given the topic’s sensitive nature, attention was given to students’ psychological comfort, as some students might have personal experiences with infertility in their family that could make certain questions emotionally triggering; it was stated that students were free to leave any uncomfortable question blank. A summary of the findings was shared with the participating institutions to benefit curriculum development, without sensitive or identifying information. Every step of the study was guided by the ethical principles of respect for persons, beneficence, and justice.

## Results

### Sociodemographic characteristics of respondents

Table 1 presents the socio-demographic characteristics of participants. A total of 265 students responded to the survey, but two did not consent to participate. A final analytic sample of 263 participants were therefore used in this report.

**Table 1.**
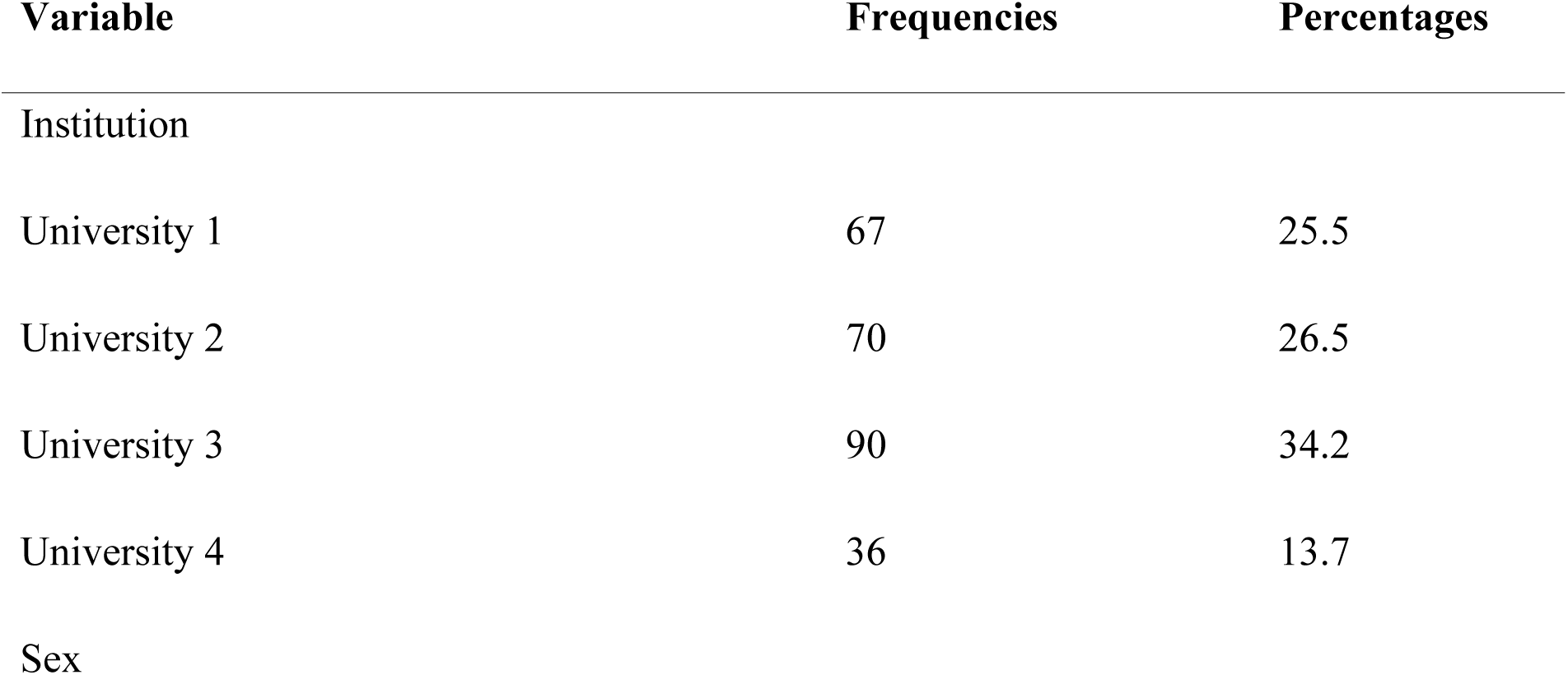
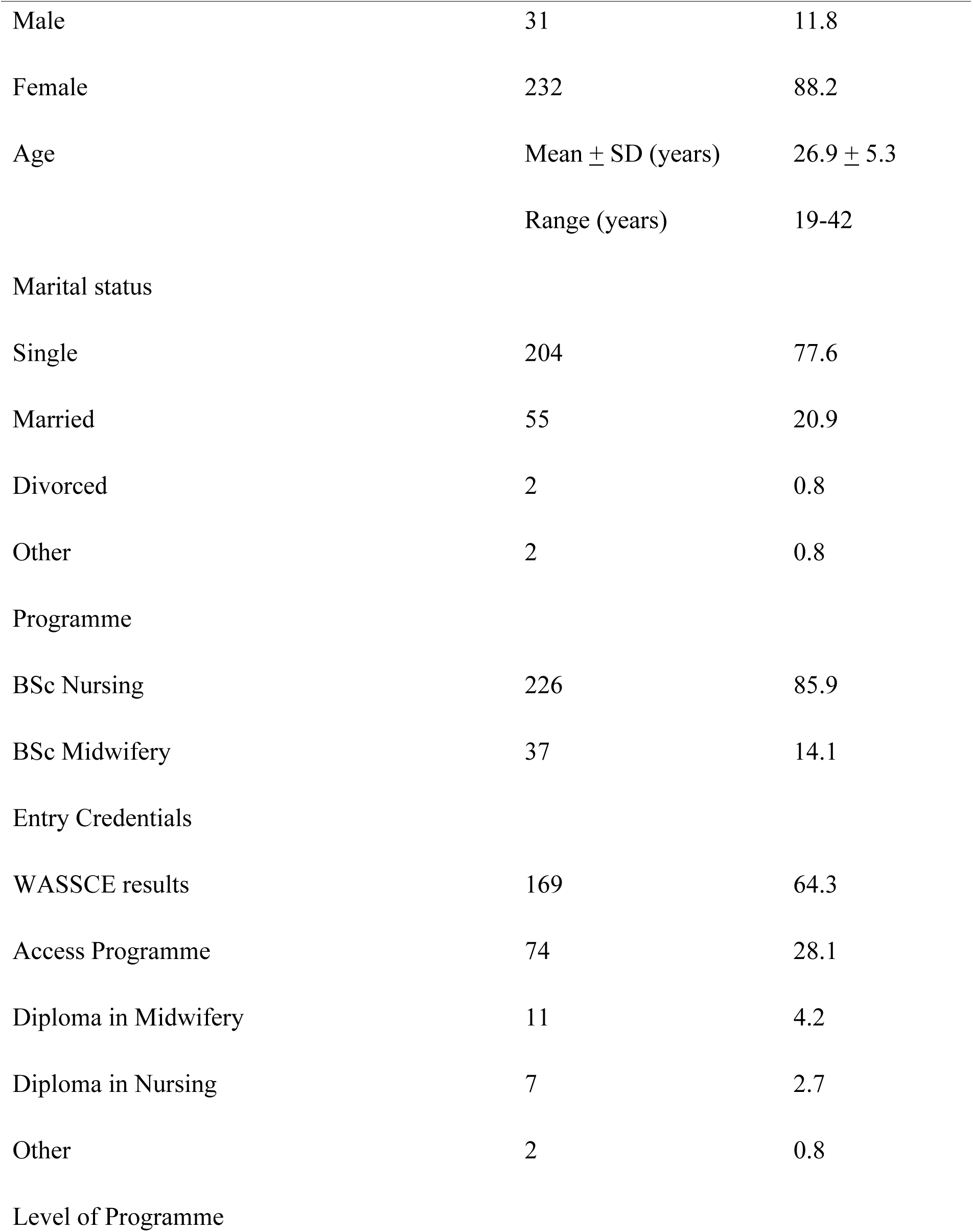
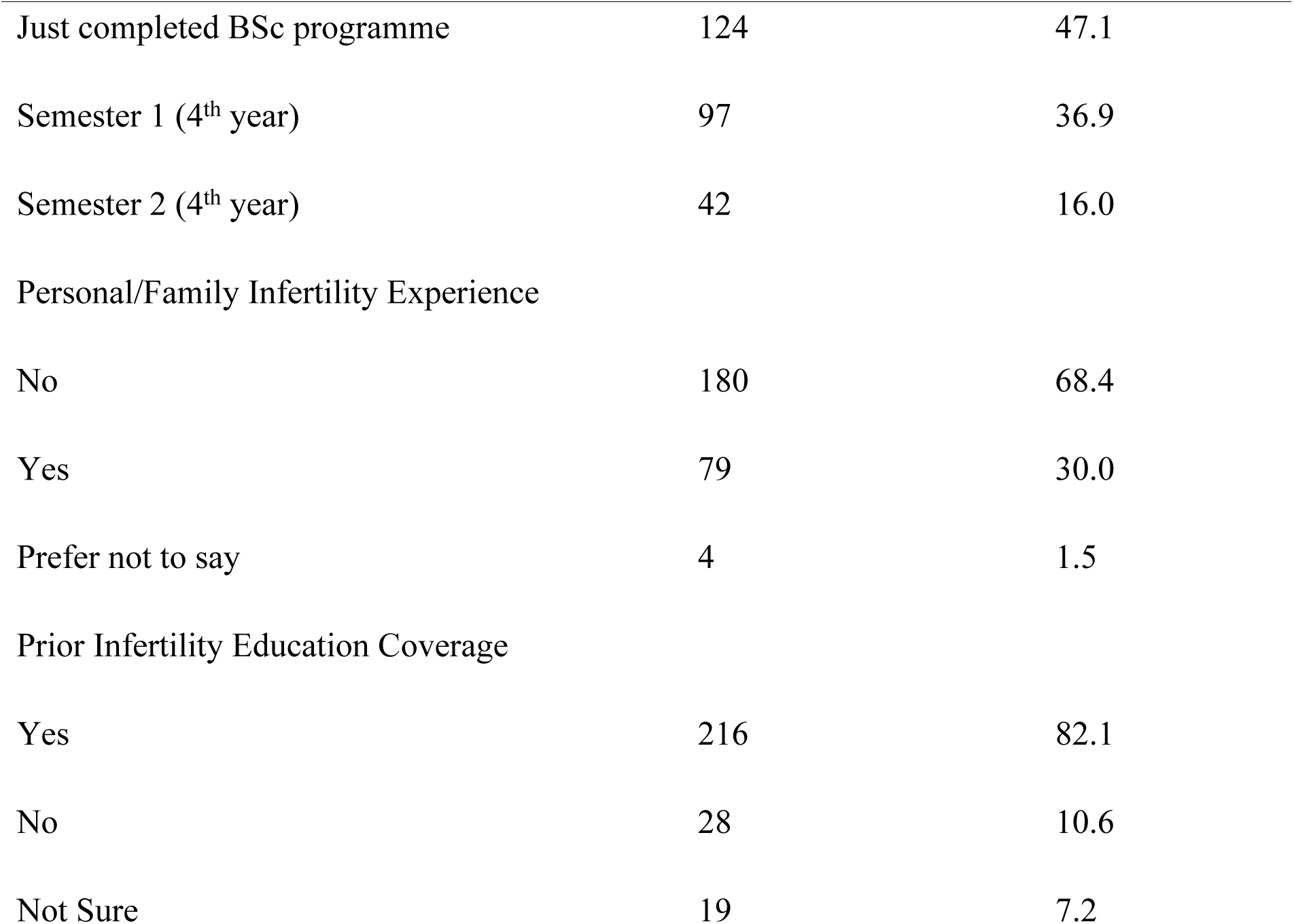
Socio-demographic characteristics of respondents.

The majority (232; 88.2%) of the participants were females, and 31 (11.8%) were males, which is broadly reflective of the demographic composition of nursing and midwifery training cohorts in Ghana. The mean age of participants was 26.9 years (SD = 5.3; range 19-42 years), with one extreme outlier (an evidently erroneous data entry) set to missing, leaving a valid age N of 262. The majority (204; 77.6%) were single, with 55 (20.9%) reporting as married.

By institution, University 3 contributed the largest (90; 34.2%) share of respondents, followed by university 2 (70; 26.6%), university 1 (67; 25.5%), and university 4 (36; 13.7%). The dominant programme of enrolment was BSc Nursing (226; 85.9%), with BSc Midwifery students comprising 37 (14.1%) of the sample; two respondents who indicated non-standard programme classifications were recoded into BSc Nursing. The most common entry qualification was the WASSCE (West African Senior School Certificate Examination (169; 64.3%), followed by Access Programme holders (i.e., certificate in nursing holders who had been upgraded) (74; 28.1%). With respect to academic progression, 124 respondents (47.1%) reported having just completed their BSc programme, while 97 (36.9%) were enrolled in Semester 1 and 42 (16.0%) in Semester 2 of their final year.

Regarding personal or familial experience with infertility, 79 (30%) respondents reported that a known family member or they personally had experienced infertility, while 180 (68.4%) reported no such experience and four (1.5%) preferred not to say anything. The substantial majority (216; 82.1%) of respondents stated that infertility had been covered as a topic in at least one course during their training, while 28 (10.6%) reported that it had not, and 19 (7.2%) were unsure.

### Knowledge of infertility

#### Item-level knowledge performance

Respondents’ knowledge of infertility was assessed across 13 scored knowledge dimensions spanning definition, aetiology, epidemiology, misconception correction, treatment recognition, and preventability. Table 2 presents the percentage of correct responses for each item or the mean proportional score for multi-select items.

**Table 2.** Item-level knowledge performance: percentage of correct responses (N=263)

| Item | Description | Correct Response | Correct / Mean Score | % Correct |
|---|---|---|---|---|
| B1 | Clinical definition of infertility | B: 12-month criterion | 217 | 82.5 |
| B2 | Infertility affects both sexes | True | 262 | 99.6 |
| B3 | Causes of infertility in women (max 4 pts) | A, B, D, F | 3.56/4 | 89.1 |
| B4 | Causes of infertility in men (max 5 pts) | A, B, C, D, E | 4.17/5 | 83.3 |
| B5 | Age-related fertility decline | True | 238 | 90.5 |
| B6 | Population prevalence of infertility | C: around 10-15%+ | 182 | 69.2 |
| B7A | Single abortion → infertility | False | 164 | 62.4 |
| B7B | Modern contraception → lasting infertility | False | 176 | 66.9 |
| B7C | Untreated STIs → infertility | True | 251 | 95.4 |
| B7D | About 50% of infertility cases involve male factors | True | 86 | 32.7 |
| B7E | 6-month criterion applies to women over 35 years | True | 97 | 36.9 |
| B8 | Recognised medical treatments for managing infertility (max 4 pts) | A, B, C, F | 3.21/4 | 80.1 |
| B9 | Infertility is at least partly preventable | B: Yes, partly | 251 | 95.4 |

With regard to the clinical definition of infertility (B1), 217 respondents (82.5%) correctly identified the 12-month criterion for regular unprotected intercourse as the standard clinical definition. Almost all respondents (n = 262; 99.6%) correctly identified that infertility can affect both men and women (B2), indicating near-universal awareness of its bilateral nature.

On the multi-select item concerning causes of infertility in women (B3), respondents achieved a mean score of 3.56 out of a possible 4 points (89.1%), reflecting strong recognition of blocked fallopian tubes, ovulatory problems, untreated sexually transmitted infections (STIs), and uterine abnormalities as valid aetiological factors. Similarly, for male causes of infertility (B4), the mean score was 4.17 out of 5 (83.3%), with most respondents identifying low sperm count, impotence, substance use, tight undergarments, and prior genital infections. Knowledge of age-related fertility decline was high, with 238 respondents (90.5%) correctly affirming that a 45-year-old woman is more likely to face conception difficulties than a 25-year-old (B5). Regarding the estimated prevalence of infertility, 182 respondents (69.2%) correctly selected the option indicating that infertility affects approximately 10-15% or more of couples (B6), though 42 (16.0%) indicated they had no idea.

On the five-item misconception sub-scale (B7A-B7E), performance was markedly more variable. Correct rejection of the belief that a single prior abortion is likely to cause infertility (B7A: False) was achieved by 164 (62.4%) respondents, and correct rejection of the claim that modern contraception causes lasting infertility (B7B: False) was demonstrated by 176 (66.9%) respondents. The near-consensus item was B7C, correctly affirming that untreated STIs can lead to infertility, which was answered correctly by 251 (95.4%) respondents. In contrast, B7D affirming that approximately half of infertility cases involve male-factor aetiology, was answered correctly (32.7%) by only 86 respondents, making it the weakest-performing item in the entire scale. Item B7E, affirming that clinical guidelines recommend a shorter diagnostic threshold of six months for women aged 35 or older, was correctly answered by 97 (36.9%) respondents, the second-lowest item performance recorded.

On the multi-select treatment item (B8), the mean score was 3.21 (80.1%) out of a possible 4 points, indicating that the majority recognised ovulation-stimulating medications, IVF, surgical correction, and intrauterine insemination as legitimate medical options. Finally, 251 (95.4%) respondents correctly identified that infertility is at least partly preventable through strategies such as treating infections early and maintaining a healthy lifestyle (B9).

#### Composite knowledge score and classification

The composite knowledge score, ranging from 0 to 23, yielded a mean of 18.25 (SD = 3.17), corresponding to a mean percentage score of 79.4%. The minimum score recorded was 6 and the maximum was 23, indicating a distribution skewed toward higher levels of knowledge. Applying the pre-specified classification thresholds, the vast majority of respondents were classified as demonstrating a high level of knowledge (n = 225; 85.6%), while 35 respondents (13.3%) fell into the moderate category and only three respondents (1.1%) were classified as low. Table 3 presents the distribution of composite knowledge scores by classification category.

**Table 3.** Distribution of composite knowledge score by classification category (N = 263)

| Knowledge category | Score range | n | % | Cumulative % |
|---|---|---|---|---|
| High | 16-23 | 225 | 85.6 | 100.0 |
| Moderate | 8-15 | 35 | 13.3 | 14.0 |
| Low | 0-7 | 3 | 1.1 | 1.1 |
| Total |  | 263 | 100.0 |  |

### Attitudes toward infertility

#### Reliability of the attitude scale

Prior to reporting the attitude findings, the internal consistency of the 16-item attitude scale was assessed. Items C4, C8, C10, C11, and C15 were reverse-coded to ensure directional consistency, such that higher scores uniformly reflect more favourable attitudes toward infertility and those affected by it. Cronbach’s alpha for the full 16-item scale was α = .850, indicating good internal consistency and confirming the scale’s reliability for use in this study.

#### Composite attitude score and classification

After reverse-coding the designated items, the composite attitude score, computed as the mean of all 16 items had a mean of 3.812 (SD = 0.654), with scores ranging from 1.75 to 4.94 on the 1-5 scale. Applying the pre-specified classification thresholds, 227 (86.3%) respondents were classified as holding favourable attitudes toward infertility, six (2.3%) fell within the neutral category, and 30 (11.4%) respondents demonstrated unfavourable attitudes. Table 4 summarises the composite attitude score distribution.

**Table 4.** Composite attitude score distribution and classification (N = 263)

| Attitude Category | Criterion | n | % |
|---|---|---|---|
| Favourable | Mean score > 3.0 | 227 | 86.3 |
| Neutral | Mean score = 3.0 | 6 | 2.3 |
| Unfavourable | Mean score < 3.0 | 30 | 11.4 |

### Group comparisons of knowledge and attitude scores

#### Differences by sex

Given the non-normal distribution of the composite knowledge score (negatively skewed) and the small male subgroup (n=31), non-parametric Mann-Whitney U tests were used for all sex-based comparisons. Female respondents demonstrated a significantly higher composite knowledge score than male respondents (Female: M = 18.41, SD = 3.03; Male: M = 17.06, SD = 3.91; U = 2800.5, p = .044), with a small effect size (rank biserial r = .221). See table 5.

**Table 5.**
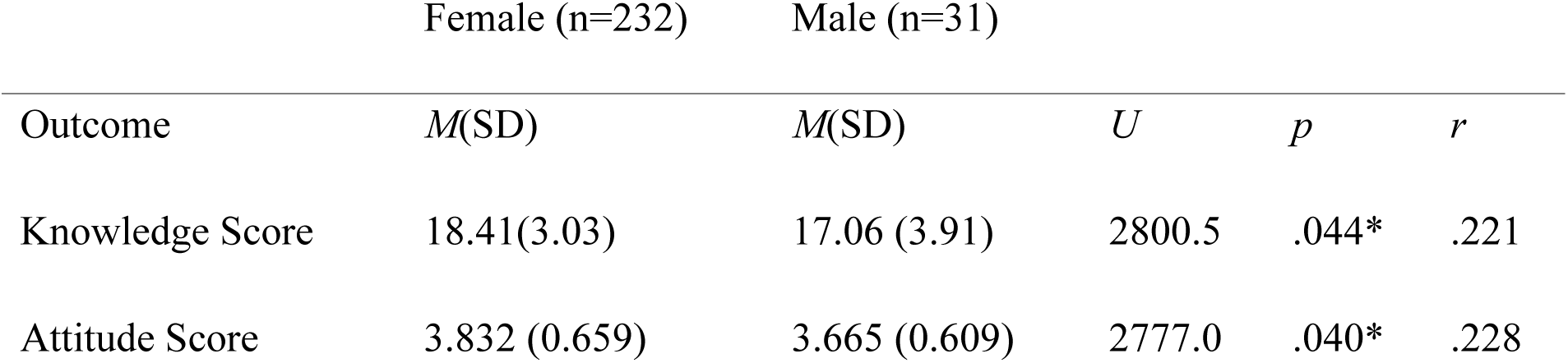
Comparison of composite knowledge and attitude scores by sex (Mann-Whitney U test).

|  | Female (n=232) | Male (n=31) |  |  |  |
|---|---|---|---|---|---|
| Outcome | *M*(SD) | *M*(SD) | *U* | *p* | *r* |
| Knowledge Score | 18.41(3.03) | 17.06 (3.91) | 2800.5 | .044* | .221 |
| Attitude Score | 3.832 (0.659) | 3.665 (0.609) | 2777.0 | .040* | .228 |

Female respondents similarly recorded a significantly higher mean composite attitude score (Female: M = 3.832, SD = 0.659; Male: M = 3.665, SD = 0.609; U = 2777.0, p = .040; r = .228). These results indicate that, while statistically significant, the magnitude of the sex differences was modest.

#### Differences by programme

A Mann-Whitney U test comparing BSc Nursing and BSc Midwifery students on composite knowledge scores revealed a statistically significant difference (BSc Midwifery: M = 19.35, SD = 2.54; BSc Nursing: M = 18.07, SD = 3.23; U = 2868.0, p = .002), with midwifery students demonstrating meaningfully higher knowledge. However, the corresponding attitude comparison did not reach statistical significance (BSc Midwifery: M = 3.876; BSc Nursing: M = 3.801; p = .502), indicating that programmatic differences in knowledge did not extend to differences in attitudinal orientation. See table six.

**Table 6.** Group comparisons of composite knowledge and attitude scores by key demographic.

| Variable | Group | n | Knowledge M(SD) | Attitude M(SD) | Test statistics | *p* |
|---|---|---|---|---|---|---|
| Sex | Female | 232 | 18.41 (3.03) | 3.832 (0.659) | U= 2800.5 / 2777.0 | .044* / .040* |
|  | Male | 31 | 17.06 (3.91) | 3.665 (0.609) |  |  |
| Programme | BSc Nursing | 226 | 18.07 (3.23) | 3.801 (0.664) | U = 2868.0 | .002** / .502 |
|  | BSc Midwifery | 37 | 19.35 (2.54) | 3.876 (0.617) |  |  |
| Institution | University 1 | 67 | 18.22 (2.80) | 3.829 (0.637) | H (3) = 0.905 / 3.155 | .824 / .368 |
|  | University 2 | 70 | 18.43 (2.73) | 3.892 (0.548) |  |  |
|  | University 3 | 90 | 18.12 (3.94) | 3.703 (0.723) |  |  |
|  | University 4 | 36 | 18.28 (2.47) | 3.898 (0.684) |  |  |
| Fam. Infertility Exp. | Yes | 79 | 18.34 (3.10) | - | U = 7285.5 | .750 |
|  | No | 180 | 18.16 (3.20) | - |  |  |
| Prior Educ. Coverage | Yes | 216 | 18.34 (3.03) | - | H(2) = 3.832 | .147 |
|  | No | 28 | 16.96 (4.02) | - |  |  |
|  | Not sure | 19 | 19.11 (2.92) | - |  |  |

### Association between knowledge and attitude scores

To assess the association between knowledge and attitude, the central relational hypothesis of the KAP framework, Pearson and Spearman correlation analyses were conducted on the composite scores. A statistically significant moderate positive correlation was observed between the composite knowledge score and the composite attitude score (Pearson r = .400, p < .001; Spearman ρ = .363, p < .001; N = 263), indicating that respondents with greater knowledge of infertility tended to hold more favourable attitudes. The consistency between parametric and non-parametric correlation estimates supports the robustness of this association. Table 7 presents the correlation results.

**Table 7.**
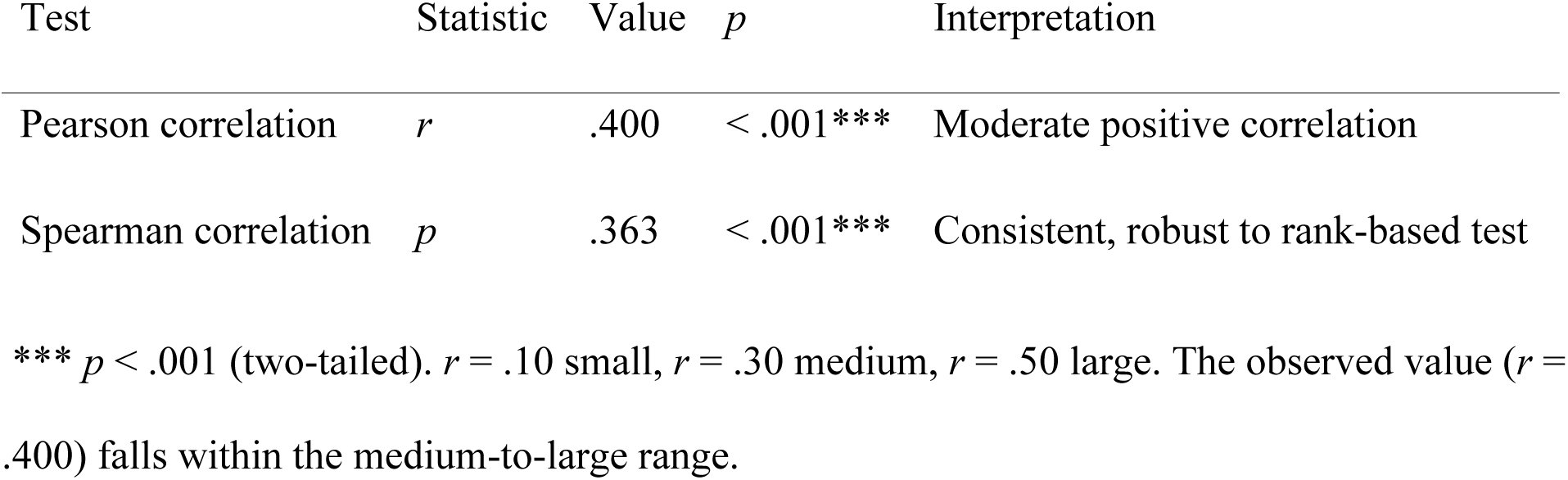
Pearson and Spearman Correlation Between Composite Knowledge Score and Composite Attitude Score (N = 263)

### Multiple linear regression: predictors of attitude score

To identify independent predictors of the composite attitude score, a multiple linear regression model was specified with the following predictors: composite knowledge score, sex (female = 1), programme (midwifery = 1), personal or familial infertility experience (yes = 1), prior infertility education coverage (yes = 1), and three institution dummy variables (reference category: University 1). The model was estimated using ordinary least-squares regression on the complete analytic sample (N = 263). Results are presented in Table 8.

**Table 8.** Multiple linear regression: predictors of composite attitude score (N = 263)

| Predictor | *B* | *SE* | *r* | *p* | 95% CI |
|---|---|---|---|---|---|
| Constant | 2.346 | 0.251 | 9.35 | < .001*** | [1.854, 2.838] |
| Knowledge Score | **0.080** | **0.012** | **6.685** | **< .001\*** | [0.057, 0.104] |
| Sex (Female = 1) | 0.062 | 0.118 | 0.519 | .604 | [−0.171, 0.294] |
| Programme (Midwifery = 1) | 0.074 | 0.111 | 0.672 | .502 | [−0.143, 0.291] |
| Family Infertility Exp. (Yes = 1) | −0.054 | 0.082 | −0.655 | .513 | [−0.214, 0.107] |
| Institution 4: University 4 | 0.056 | 0.125 | 0.447 | .655 | [−0.189, 0.301] |
| Institution 2: University 2 | 0.068 | 0.105 | 0.644 | .520 | [−0.139, 0.274] |
| Institution 3: University 3 | −0.121 | 0.099 | −1.226 | 221 | [−0.314, 0.072] |

The overall model was statistically significant (F(8, 254) = 6.851, p < .001), accounting for 17.8% of the variance in attitude scores (R² = .178, adjusted R² = .152). Inspection of individual predictors revealed that the composite knowledge score was the sole statistically significant predictor of attitude (B = 0.080, SE = 0.012, t(254) = 6.685, p < .001), indicating that for each one-point increase in the composite knowledge score, the attitude score increased by approximately 0.08 points after controlling for all other variables. No other predictor reached statistical significance, including sex (B = 0.062, p = .604), programme (B = 0.074, p = .502), familial infertility experience (B = −0.054, p = .513), prior infertility education (B = −0.038, p = .709), or institutional affiliation. Variance inflation factors (VIF) for all predictors ranged from 1.017 to 1.584, well below the commonly applied threshold of 5.0, confirming the absence of multicollinearity.

### Practice and behavioural intentions

With regard to clinical exposure, 65 respondents (24.7%) reported having encountered at least one infertility case during clinical rotations or community health postings, while 198 (75.3%) had not. Those who reported clinical exposure provided brief accounts of their responses, typically describing referral of clients to specialists, provision of emotional support, or documentation of the case as an observation. Table 9 summarises practice and behavioural intention responses.

**Table 9.** Descriptive summary of practice and behavioural intention responses (N = 263)

| Item | Response option | n | % |
|---|---|---|---|
| D1: Clinical exposure to infertility | Yes, encountered cases | 65 | 24.7 |
|  | No | 198 | 75.3 |
| D2: Likelihood to initiate fertility conversation | Very likely | 127 | 48.3 |
|  | Likely | 101 | 38.4 |
|  | Not sure | 101 | 10.3 |
|  | Unlikely | 7 | 2.7 |
|  | Very unlikely | 1 | 0.4 |
| D3: Should curriculum include more infertility content? | Yes, definitely | 227 | 86.3 |
|  | Maybe | 31 | 11.8 |
|  | No, current content is enough | 3 | 1.1 |
|  | I'm not sure | 2 | 0.8 |
| D4: Interest in infertility workshop or short course | Yes | 238 | 90.5 |
|  | Maybe | 25 | 9.5 |
|  | No | 0 | 0.0 |

Regarding future clinical intentions, 127 respondents (48.3%) indicated they would be very likely to initiate conversation about fertility with clients in a family planning or antenatal setting, and a further 101 (38.4%) indicated they were likely to do so, yielding a combined proportion of 86.7% indicating at least some readiness to engage proactively. Only eight respondents (3.0%) expressed unlikely or very unlikely disposition to initiate such conversations.

The vast majority of respondents supported enhanced infertility content in nursing and midwifery training: 227 respondents (86.3%) agreed that the curriculum should include more content on fertility and infertility, while 31 (11.8%) selected “maybe.” Only five respondents (1.9%) indicated that the current level of content was sufficient or that they were unsure. Interest in a future continuing education workshop or short course on infertility management was similarly high, with 238 respondents (90.5%) expressing definite interest and the remaining 25 (9.5%) responding “maybe.” Not a single respondent declined interest in such a workshop

### Qualitative summary of open-ended responses

Approximately 60 respondents provided written comments in response to the optional open-ended item D5, which invited additional thoughts on infertility or the nursing role. Thematic content analysis of these responses identified five dominant themes.

The most frequently expressed theme was **curriculum enhancement**, with multiple respondents calling for greater depth and breadth of infertility content in nursing and midwifery training, such as “Nurses and midwives should be given more education and training on how to deal with infertility issues” and “Nursing education and the content of fertility and infertility should be spread widely.” A closely related theme concerned **clinical training and workshop needs**, with respondents specifically requesting practical, skills-based training formats: “Workshops should be held for nurses and midwives to help them know more about modern care for infertility” and “Training on how to support couples with fertility issues.”

The third prominent theme addressed the need for **non-judgmental and empathic care**, reflecting respondents’ awareness that negative nursing attitudes exist and represent a barrier to care: “The attitudes of some nurses and the way we are very judgemental when it comes to sensitive topics is nothing to write home about” and “Nurses should be more loving, confidential, and non-judgemental so clients can come to us when the need arises.”

The fourth theme concerned the nurses’ **community education and destigmatisation** role: “More education should be done to make the public more aware and to reduce stigma associated with infertility” and “Nurses can help reduce infertility stigma by offering accurate education, emotional support, and early referrals for care.” Finally, a fifth theme of **psychosocial support and counselling** was expressed by respondents who emphasised the emotional dimensions of infertility: “Couples with infertility are going through a lot, so we must treat them with respect” and “Infertility can have a deep emotional, psychological, and social impact on individuals and couples.”

## Discussion

### Knowledge and attitudes toward infertility

This study examined the relationship between knowledge and attitudes toward infertility and reproductive health among final-year nursing and midwifery students in Accra, Ghana. The composite knowledge score (M = 18.25/23, 79.4%) indicates a high overall baseline of factual knowledge, with 85.6% of respondents classified in the “High” knowledge category. This finding diverges from the broader regional literature, in which infertility knowledge among tertiary students and even practicing health professionals has generally been reported as limited. A research study [11] for instance, found that knowledge about infertility was quite limited among Ghanaian university students outside nursing and medical cohorts, and another study [17] found that over half of Turkish nursing students believed infertility “cannot be treated.” The comparatively strong performance of the present sample may reflect the fact that 82.1% of respondents reported having covered infertility as a topic during their training, suggesting that exposure to reproductive health content within the nursing and midwifery curriculum, even where infertility receives only minimal dedicated coverage, as the literature review notes, may be sufficient to produce adequate factual recall on core definitional and aetiological items.

However, the item-level analysis revealed two notable and clinically consequential gaps that the aggregate score conceals. Only 32.7% of respondents correctly identified that approximately half of infertility cases involve male-factor aetiology, and only 36.9% correctly identified the shortened six-month diagnostic threshold recommended for women over 35 years. The first gap is consistent with the broader pattern, documented extensively in the literature review, of male-factor infertility being underemphasized and under-discussed in both lay and clinical discourse in sub-Saharan Africa, with blame and social repercussions disproportionately falling on women [3, 5]. That nearly two-thirds of final-year nursing and midwifery students, who are on the cusp of independent practice, underestimate the male contribution to infertility suggests that this sociocultural bias may be reproduced rather than corrected within formal training, with direct implications for how future providers counsel couples and allocate diagnostic attention. The second gap, concerning the age-adjusted diagnostic window, reflects a more technical clinical knowledge deficit that has direct bearing on the timeliness of referral and management decisions for women approaching the end of their reproductive years.

Attitudes toward infertility were predominantly favourable (M = 3.812/5; 86.3% classified “Favourable”), with particularly strong endorsement of the professional and empathic role of nurses and midwives in supporting infertile couples, and correspondingly low endorsement of stigmatising beliefs such as infertility being a punishment or spiritual sign. This pattern echoes, and arguably exceeds, the moderately positive attitudes among Turkish nursing students [17]. It suggests that, despite the persistence of stigmatising sociocultural narratives in the wider Ghanaian context documented throughout the literature review [2, 9, 19], formal nursing and midwifery education in Accra may be functioning as a partial counterweight to those narratives, equipping students with a professionally oriented, biomedically grounded outlook even where some underlying knowledge gaps persist.

### Association between knowledge and attitude

A statistically significant moderate positive correlation was found between composite knowledge and attitude scores (r = .400, p < .001), and knowledge score emerged as the sole significant predictor of attitude in the multiple regression model (B = 0.080, p < .001), after accounting for sex, programme, institution, family infertility experience, and prior infertility education coverage. This finding provides empirical support for the central tenet of the Knowledge-Attitude-Practice (KAP) model underpinning this study, namely that knowledge acquisition is theorized to shape attitudes, which in turn are expected to influence practice [13]. The fact that knowledge remained significant while none of the demographic or institutional variables did suggests that, within this population, cognitive mastery of the subject matter is a more proximal driver of favourable orientation than background characteristics or where a student trained. This is a reassuring finding for curriculum designers: it implies that closing specific knowledge gaps, such as those identified for male-factor aetiology and age-related diagnostic criteria, is a tractable, modifiable lever through which more favourable attitudes might be further reinforced, rather than attitudes being fixed by less malleable factors such as institutional culture or personal background.

At the same time, the regression model explained only 17.8% of the variance in attitude scores, indicating that the majority of variation in students’ attitudes is attributable to factors not captured in this study, such as personality, prior exposure to infertile individuals beyond the binary “family experience” variable measured here, religiosity, or broader empathy disposition. This is consistent with the KAP literature’s caution that the knowledge-to-attitude linkage, while real, is neither deterministic nor sufficient on its own [15].

### Group differences by sex, programme, and institution

Female respondents demonstrated significantly higher knowledge (U = 2800.5, p = .044) and attitude scores (U = 2777.0, p = .040) than male respondents, although the effect sizes were small (r = .221 and .228, respectively) and the male subgroup was considerably smaller (n = 31) than the female subgroup (n = 232), warranting caution in generalizing this disparity. This finding is broadly consistent with earlier study which reported that male nursing students in Turkey displayed more negative attitudes and lower empathy toward infertile individuals than their female counterparts [17], and a very recent study [11] found Ghanaian female university students more likely to hold positive attitudes toward infertility issues than male students. Taken together with the present findings, this cross-cultural consistency suggests a durable pattern, rather than a context-specific artefact, and points to a need for targeted attitudinal and knowledge-reinforcement strategies aimed specifically at male nursing and midwifery trainees, who may otherwise carry comparatively under-formed competencies into a clinical workforce in which they will be expected to counsel both male and female patients on fertility matters.

BSc Midwifery students demonstrated significantly higher knowledge scores than BSc Nursing students (U = 2868.0, p = .002), though the corresponding difference in attitude scores did not reach statistical significance (p = .502). This pattern is plausible given that midwifery curricula in Ghana typically involve more concentrated and applied exposure to reproductive and maternal health content than general nursing curricula, which may translate into stronger factual recall on infertility-specific items without necessarily producing a parallel difference in general attitudinal orientation, since favourable attitudes toward infertile patients may be cultivated through broader professional socialisation common to both programmes rather than through programme-specific content alone.

There were no statistically significant differences in knowledge or attitude scores that were observed across the four participating institutions, despite their differing ownership structures (public versus private), religious affiliations, and institutional histories, as detailed in the methodology. This absence of institutional variation is a noteworthy finding in its own right: it suggests a degree of national consistency in how infertility-related content is delivered across Ghana’s nursing and midwifery training landscape, which may reflect shared adherence to Nursing and Midwifery Council curricular standards across institutions of varying type. It also strengthens the generalisability of the present findings beyond any single institutional context. Similarly, neither personal or familial infertility experience nor self-reported prior coverage of infertility in coursework was significantly associated with knowledge scores, suggesting that direct personal exposure to infertility, in the absence of structured educational content, does not by itself confer the technical knowledge gaps identified in this study; experiential familiarity and formal knowledge acquisition appear to operate through largely independent pathways.

### Practice intentions and the demand for further training

Findings from the question on practice and behavioural intentions indicate that final-year students hold strong behavioural intentions consistent with their favourable attitudes: 86.7% of respondents indicated they would be likely or very likely to initiate conversations about fertility with clients in future antenatal or family planning settings, despite only 24.7% having encountered an actual infertility case during their clinical rotations to date. This gap between limited direct clinical exposure and strong intended engagement suggests that students’ favourable attitudes and intentions are, at this stage, largely anticipatory rather than experience-tested, reinforcing the importance of structured curricular and clinical-placement interventions, rather than incidental clinical exposure alone, in preparing students for this aspect of practice. The near-unanimous demand for expanded curriculum content (86.3%) and interest in continuing education on infertility management (90.5%) reinforces this picture and aligns closely with the qualitative themes that emerged from the open-ended responses, namely calls for curriculum enhancement, clinical training, non-judgmental care, community destigmatisation, and psychosocial support, all of which converge on a consistent message that students perceive their current preparation as a foundation requiring further reinforcement rather than as already sufficient.

## Conclusion

This study found that final-year nursing and midwifery students across four universities in Accra, Ghana, possessed a generally high level of knowledge and held predominantly favourable attitudes toward infertility and the individuals affected by it. Nonetheless, specific and clinically meaningful knowledge gaps persisted regarding the proportion of infertility cases attributable to male-factor causes and the age-adjusted diagnostic criteria for women over 35 years, gaps that mirror long-documented patterns of gendered blame attribution in the wider Ghanaian sociocultural context. Knowledge and attitude were significantly and positively correlated, and knowledge score was the only significant predictor of attitude among the demographic and institutional variables examined, lending empirical support to the Knowledge-Attitude-Practice framework that guided this study. Female students and BSc Midwifery students demonstrated stronger performance on selected outcomes, while no significant differences emerged across the four participating institutions, suggesting a degree of national consistency in curricular delivery. Students expressed strong demand for expanded infertility content and continuing professional education, alongside clear behavioural intentions to engage proactively with fertility-related conversations in future practice. These findings support targeted, content-specific curricular reinforcement, particularly around male-factor infertility and age-related diagnostic thresholds, and underscore the value of deliberately structured clinical exposure to infertility care, as concrete, evidence-based steps toward strengthening reproductive health education and reducing infertility-related stigma within Ghana’s nursing and midwifery training pipeline.

## Data Availability

N/A

## Acknowledgements

The authors wish to acknowledge the authorities of the four universities who granted permission to have access to the students; and to all the Level 400 nursing and midwifery students who voluntarily participated in the study, we are most grateful.

## Author contributions

Conceptualisation: Ernestina Safoa Donkor

Methodology: Ernestina Safoa Donkor, Emmanuel Kojo Osafo. Ezekiel Oti-Boadi, Benedicta Twum-Dei

Data curation: Ernestina Safoa Donkor, Emmanuel Kojo Osafo. Ezekiel Oti-Boadi, Benedicta Twum-Dei

Formal analysis: Emmanuel Kojo Osafo, Ernestina Safoa Donkor

Writing - original draft: Ernestina Safoa Donkor

Writing - review and editing: Emmanuel Kojo Osafo, Ezekiel Oti-Boadi, Benedicta Twum-Dei,

